# Survival outcomes are associated with genomic instability in luminal breast cancers

**DOI:** 10.1101/2020.02.25.20027920

**Authors:** Lydia King, Andrew Flaus, Aaron Golden

## Abstract

Breast cancer is the leading cause of cancer related death among women. Breast cancers are generally diagnosed and treated based on clinical and histopathological features, along with subtype classification determined by the Prosigna Breast Cancer Prognostic Gene Signature Assay (also known as PAM50). Currently the copy number alteration (CNA) landscape of the tumour is not considered. We set out to examine the role of genomic instability (GI) in breast cancer survival since CNAs reflect GI and correlate with survival in other cancers. We focussed on the 70% of breast cancers classified as luminal and carried out a comprehensive survival and association analysis using Molecular Taxonomy of Breast Cancer International Consortium (METABRIC) data to determine whether CNA burden quartiles derived from absolute CNA counts are associated with survival. Luminal A and B patients were stratified by PAM50 subtype and tumour grade and then tested for association with CNA burden using multiple statistical tests. Analysis revealed that patients diagnosed with luminal A grade 3 breast cancer have a CNA landscape associated with disease specific survival, suggesting that these patients could be classified as at-risk. Furthermore, luminal A grade 3 cases largely occupy a region of stratification based on gene expression at the boundary where luminal A and luminal B cases overlap. We conclude that GI reflected by absolute CNA score is a statistically robust prognostic factor for survival in luminal A grade 3 breast cancer. Therefore, luminal A grade 3 breast cancer patients in CNA burden quartiles 3 or 4 may benefit from more aggressive therapy. This demonstrates how individual genomic landscapes can facilitate personalisation of therapeutic interventions to optimise survival outcomes.

## Introduction

Breast cancer is one of the most common malignancies affecting women worldwide and is the leading cause of cancer related death among this group [1–3]. Over 2 million new breast cancer cases were reported in 2018 and it is estimated that over 40,000 people will die as a result of breast cancer in the United States in 2019 [2, 4].

Breast cancer was previously treated as a single disease, so patients diagnosed with different histological subtypes often underwent similar treatment strategies [5].

Advances in areas such as next generation sequencing have now led to breast cancer being regarded as a collection of highly heterogeneous diseases with distinct molecular and clinical phenotypes including disease progression rate, treatment response and survival [1]. The molecular classification of breast cancer currently makes use of PAM50 intrinsic subtyping determined by the Prosigna Breast Cancer Prognostic Gene Signature Assay (formerly called the PAM50 test) [6] based on gene expression profiling [7, 8]. This distinguishes luminal A (lumA), luminal B (lumB), human epidermal growth factor receptor 2 (*HER2*)-enriched and basal-like subtypes [6]. The differences in gene expression patterns among these intrinsic subtypes reflect basic alterations in the cell biology of the tumours [9]. Importantly, it has been observed that *∼*85% of the variations in gene expression patterns of breast cancers are as a result of CNAs [1, 10].

Approximately 70% of breast cancers belong to the luminal subtypes lumA and lumB [11] characterised by increased levels of estrogen receptor (ER) and progesterone receptor (PR). LumA tumours display lower levels of genomic instability (GI) compared to lumB tumours [11]. GI is regarded as a hallmark of cancer [12] and refers to an increased tendency toward alterations in the genome during the life of cells. These alterations range from single nucleotide alterations to large scale structural changes of chromosomes, aneuploidy and whole genome duplications [12]. GI has the ability to initiate cancer, affect progression and influence patient prognosis [13].

Recent studies suggest that the relationship between lumA and lumB may be a continuum rather than a strict division of subtypes [9–11]. It has also been hypothesised that lumA tumours may evolve into lumB tumours as a result of stochastic acquisitions of mutations in genes associated with worse prognosis, including *HER2* and tumour protein p53 (*TP53*) [11, 14].

At present breast cancer diagnosis and treatment follows an integrative approach whereby both clinical and histopathological features such as age at diagnosis, tumour size, lymph node metastasis and tumour grade are utilised alongside tissue derived biomarkers [15]. However, it is widely accepted that breast cancer is largely dominated by chromosomal rearrangements [1], and a growing body of evidence suggests that the incorporation of the genomic landscape of the tumour into treatment decisions is extremely beneficial to the patient [16, 17].

Several studies have shown that the copy number landscape of a tumour can affect survival [1, 18, 19]. A pan-cancer analysis suggests that the CNA burden measured as the percentage of the tumour genome with CNAs is associated with both overall survival (OS) and disease specific survival (DSS) in a range of cancers including breast, endometrial, renal, thyroid, and colorectal cancer [19]. Assessing aneuploidy in prostate cancers at diagnosis has been shown to be more predictive of long term survival than the Gleason score which is the standard clinical metric [18]. Consistent with this, Zhang et al. [1] reported an association between general CNA burden and breast cancer survival in the Molecular Taxonomy of Breast Cancer International Consortium (METABRIC) dataset.

These studies support the conjecture that the CNA landscape of a tumour is itself associated with both OS and DSS, and could provide a prognostic biomarker [1, 18, 19]. The original association in breast cancer reported by Zhang et al. [1] considered all PAM50 intrinsic subtypes and used a simple binary measure of CNA burden to categorise GI. Tishchenko et al. (2016) used a cytoband-based measure of CNA gain/loss rate ranging from −1 to +1 correlated to local gene expression to quantify CNA variation in their analysis [11].

We hypothesised that using a more nuanced measure of CNA burden the impact of GI on luminal breast cancer survival could provide additional prognostic information, and potentially shed light on the boundary between the lumA and lumB subtypes.

## Materials and methods

A CNA score was developed using the absolute CNA profiles of all luminal patients profiled within the METABRIC consortium. This was calculated by summing the absolute value of the scores for all genes. Cases were then assigned to ranked quartiles as a first-order means of segmenting the CNA scores for analysis (Figure 1). Survival analysis was carried out for these quartiles using associated clinical data to determine survival associated variables. Statistical association tests were then applied to validate that the association between a given CNA score quartile and its survival outcomes was due to the CNA score quartile and not to a confounding variable. Finally, Cox models and the associated assumption tests were used to confirm that the survival outcomes are associated with GI in specific cohorts of luminal breast cancers. In addition, the quantile classification based on the gene expression analysis of Tishschenko et al. (2016) [11] was utilised to examine the luminal stratification associated with cases where GI affects survival. All analysis was conducted using the R statistical processing environment, and an R Shiny App was subsequently developed to expedite this work (manuscript in preparation.)

**Fig 1.**
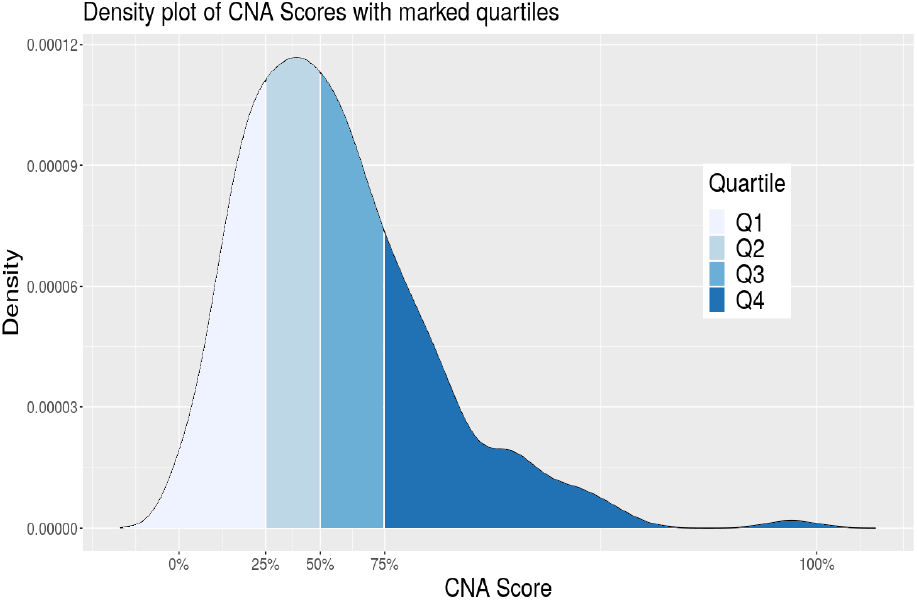
Density plot of CNA score distribution for METABRIC luminal cases. The CNA quartiles are marked out based on differing shades of blue displayed in the legend.

### METABRIC Data

METABRIC provides a well-annotated dataset of over 2,000 breast cancer cases with long-term clinical follow-up data, transcriptomic and genomic data [10]. Cases have an average follow-up time of 125 months and a maximum follow-up time of 355 months. All CNA profiles, clinical patient and sample annotations for luminal patients (n = 1175) were obtained from cBioportal [20]. The METABRIC consortium [10] utilised both the circular binary segmentation algorithm [21] and an adapted Hidden Markov model [22] for segmentation, followed by CNA calling. The patient-specific somatic CNA profile calls for each gene have values indicating homozygous deletion (−2), hemizygous deletion (−1), diploidy (0), single copy gain (+1) and high level amplification (+2). Quantile classification based on relative gene expression for luminal METABRIC cases was obtained from Tishchenko et al. [11].

### Statistical Analyses

Clinical data and CNA profiles were extracted using Python (version 3.6.3) and analysed using R (version 3.5.1) and RStudio (version 1.2.1335) with R packages *survival, survminer* and *gglplot2* [23–25]. Additional functions such as mutation analysis using the R package *maftools* [26] were also implemented. These packages and associated processing scripts were packaged into a bespoke R Shiny app with multiple tab panels capable of running and displaying the results of the entire statistical analyses (Figure 2). Sidebar tabs include Input Files, Exploratory Tables, CNA Score Distribution, Survival Analysis, Cox Regression, Maftools Plots, Density Plots and Calculations. The app provided a rapid, powerful and effective means to explore, segment, visualise and statistically test the METABRIC data.

**Fig 2.**
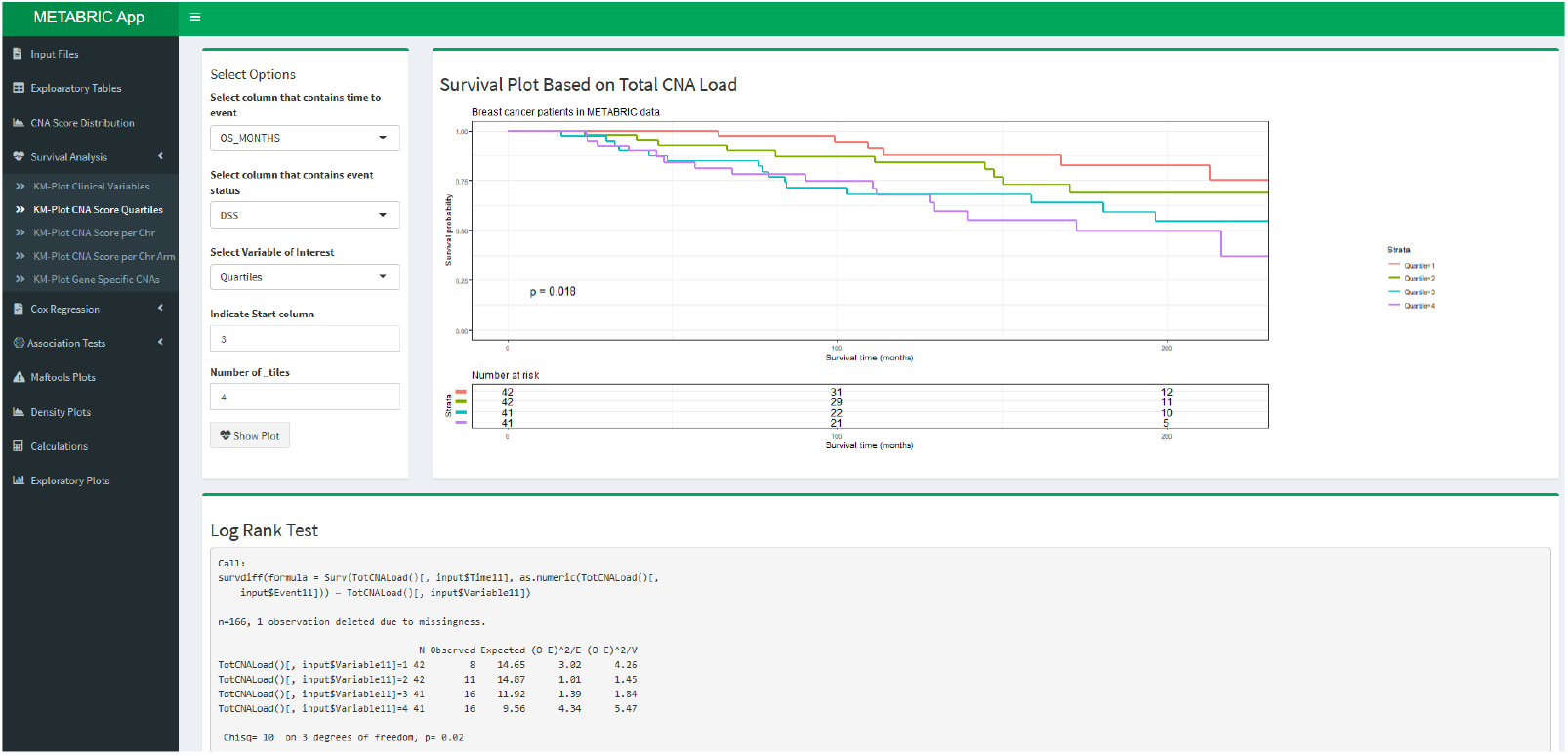
Graphical user interface (GUI) of the R shiny app (manuscript in preparation). Displayed here is the Survival Analysis tab showing the DSS based on CNA quartiles in lumA grade 3. Results from the corresponding log-rank test are displayed in the bottom box.

## Results

### Survival Outcome Is Associated With CNA Quartile

#### Survival Analysis of Luminal Breast Cancers

A number of recent studies report that CNAs reflecting GI are associated with survival outcomes in several types of cancer [1, 18, 19]. We hypothesised that CNA quartiles based on absolute CNA score would be associated with both OS and DSS in luminal breast cancer patients. Patients within quartile 4 (Q4) have higher CNA scores indicative of higher levels of GI and significantly worse survival outcomes than patients in quartiles 1-3 (Q1-3) with less GI (Figure 3, *p*-values < 0.0001). Patients in Q4 had OS and DSS rates of 32% and 61% respectively, while patients in Q1 had OS and DSS rates of 48% and 82% respectively.

**Fig 3.**
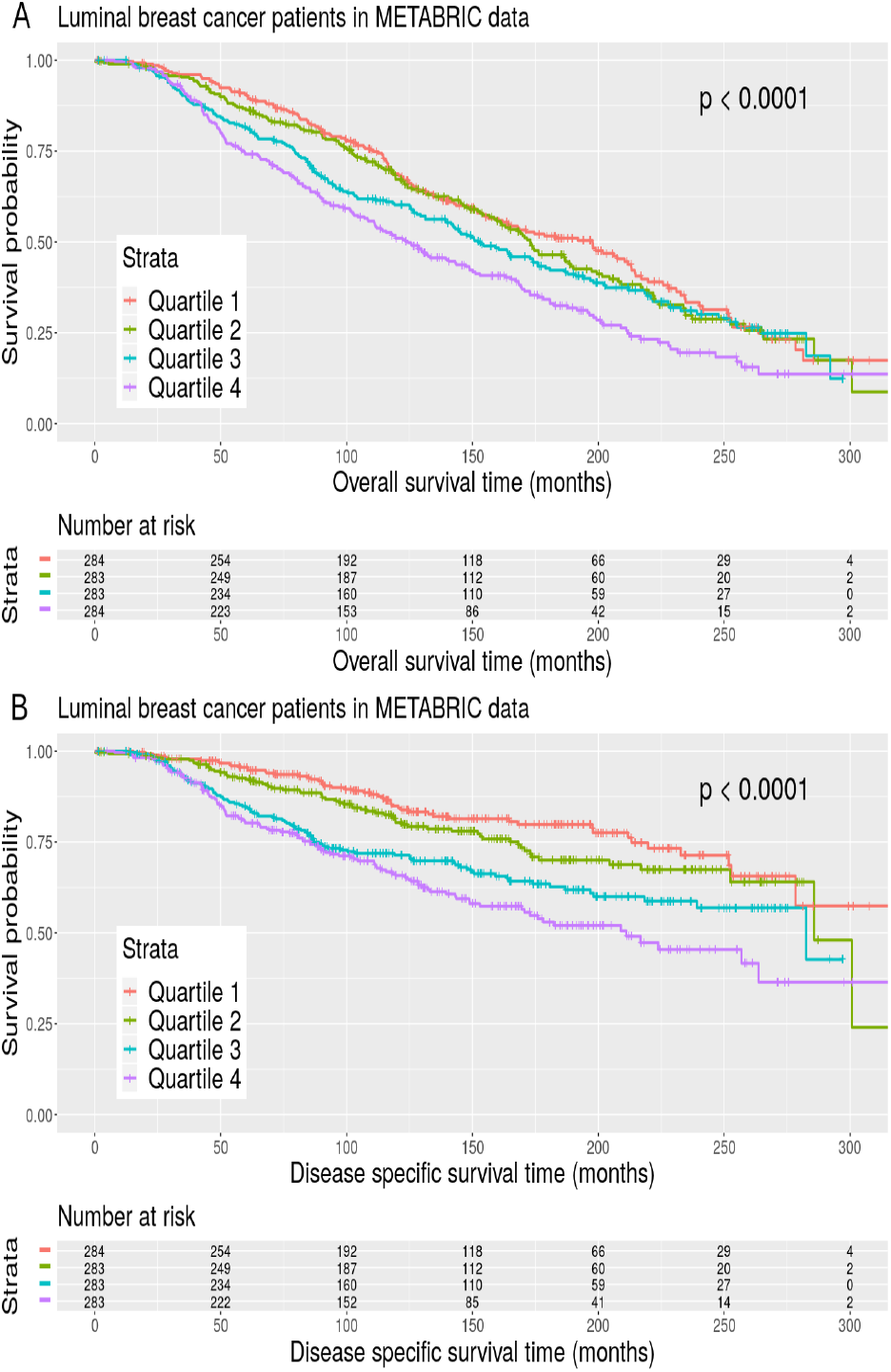
OS and DSS according to CNA quartiles in METABRIC luminal breast cancer patients. KM plots for (A) OS for CNA quartiles in METABRIC luminal data. (B) DSS for CNA quartiles in METABRIC luminal data. The *p*-value associated with the log-rank test and a risk table displaying the number of patients at risk at each time interval is displayed.

#### Possible Confounding Variables

A strong association between clinical variables such as PAM50 subtype, tumour stage and age at diagnosis and breast cancer survival has been reported [27–29]. A number of steps were taken to determine whether the association between survival outcomes and CNA quartiles was the result of confounding variables, which are additional factors influencing survival outcomes that are correlated with CNA quartiles.

First, a survival analysis using Kaplan-Meier (KM) plots and univariate Cox models was carried out to identify whether any of 23 clinical variables (Supplementary Table 1) and CNA quartiles in the luminal data were associated with survival outcome. We found that 20 of the former and the CNA quartiles were associated with either OS or DSS, or both, (Supplementary Table 3) and these were taken forward for examination using statistical tests. A *χ*^2^ test was used to test the association between two categorical variables with sufficient cell sizes in the two-way table of categorical variables. Fisher’s exact test was used in the case where any cell size was sufficiently small. The non-parametric Kruskal-Wallis test was used to determine if there were statistically significant differences between CNA quartiles and continuous clinical variables associated with survival outcomes. These tests indicated that the CNA quartiles are significantly associated with a number of clinical variables (Supplementary Table 4).

Nottingham prognostic index (NPI), PAM50 subtype and grade were three of the most significant confounding factors. Multivariate Cox regression models were used to examine the survival association and assumptions of these factors. The results indicated that all the variables examined are significantly associated with OS or DSS, or both, and the overall *p*-value indicates that the model is statistically significant (Supplementary Table 5). The assumptions of the multivariate Cox regression model for OS and DSS were tested and showed that the test was not statistically significant for any of the covariates except CNA quartile 3 (Q3) in both OS and DSS, and that the global test is also not statistically significant for DSS (Supplementary Figure 2-3). Therefore, proportional hazards (PH) can be assumed for the clinical variables and it can be concluded that NPI, PAM50 subtype and grade significantly affect survival and cannot be adjusted for in the model. This result confirms that the association between CNA quartiles and survival outcomes are a consequence of confounding variables.

### Survival Analysis Of Subsets Can Isolate Confounding Variables

#### Separate Analysis of lumA and lumB Cases

The results of the univariate Cox analyses showed that the relationship between survival and PAM50 subtype was highly statistically significant (Supplementary Table 3). Furthermore, as the CNA quartiles progress from Q1 to Q4 so too does the proportion of lumB tumours (Supplementary Figure 1). This is expected as lumA tumours generally display lower levels of GI than lumB tumours [11].

To remove the confounding effect of the PAM50 intrinsic subtype, the lumA and lumB cases were segregated and the analyses repeated (Supplementary Results) with each group again assigned to subtype CNA quartiles (Supplementary Figures 4 and 9). The subtype groups were found to be significantly associated with OS and DSS for both lumA (Supplementary Figures 5 and 6, and Supplementary Table 6) and lumB (Supplementary Figures 10 and 11, and Supplementary Table 12). However, a number of confounding factors were again identified. For lumA cases the most significant confounding variables were NPI, three gene classification, integrative cluster and grade, while for lumB the most significant confounding variables were NPI, three gene classification, integrative cluster and HER2 status (Supplementary Tables 7 and 13). These confounding variables could not be adjusted for in our survival models (Supplementary Results) so further stratification was necessary.

#### Analysis of lumA and lumB Cases by Grade

Analysis of lumA and lumB cases revealed that NPI was highly associated with both survival and the CNA quartiles, so NPI can be considered as a confounding factor. NPI incorporates tumour grade so the lumA and lumB cases were further stratified by grade. CNA quartiles were not associated with OS or DSS in lumA grade 1 and 2 cases and lumB grade 2 and 3 cases (Supplementary Figures 14-19 and 26-31), or OS in lumA grade 3 and lumB grade 1 cases (Figure 4A and Supplementary Figure 24). However, significant association between CNA quartiles and DSS was observed in lumA grade 3 and lumB grade 1 patients (Figure 4B and Supplementary Figure 25, *p*-value = 0.018 and 0.046 respectively). The association for lumB grade 1 patients was viewed with caution due to the small sample size per quartile.

**Fig 4.**
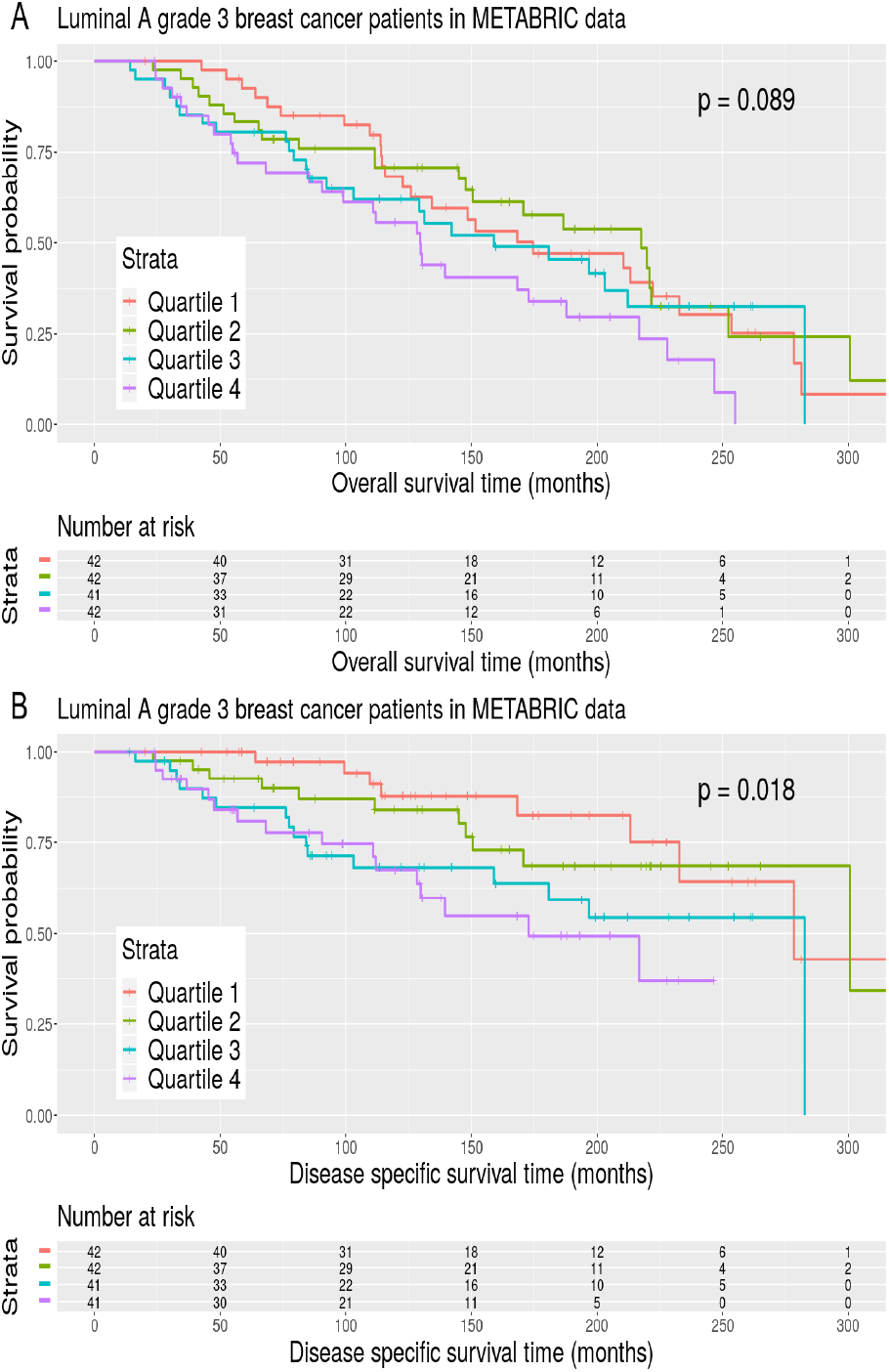
OS and DSS according to CNA quartiles in lumA grade 3 METABRIC breast cancer patients. KM plots for (A) OS for CNA quartiles in lumA grade 3 METABRIC data. (B) DSS for CNA quartiles in lumA grade 3 METABRIC data. The *p*-value associated with the log-rank test and a risk table displaying the number of patients at risk at each time interval is displayed.

#### LumA grade 3 survival is Associated with GI

To confirm that the observed association of CNA quartiles and DSS for lumA grade 3 cases was the result of a true association, a univariate Cox and KM analysis was carried out using a selection of relevant clinical characteristics that had an association with OS, DSS, or both, including the number of positive lymph nodes, NPI, estrogen receptor immunohistochemistry (ER IHC), inferred menopausal state, PR status, radiotherapy, breast surgery, tumour stage, tumour size, age at diagnosis and CNA quartiles (Supplementary Table 16). Statistical tests were used to determine which of these variables associated with survival are also associated with the CNA quartiles and thus could be potential confounding variables (Supplementary Table 17). Only age at diagnosis was significantly associated with the CNA quartiles meaning it was a confounding factor in the OS analysis.

To formally quantify the effects of CNA quartiles on OS and DSS in lumA grade 3 patients, Cox PH models were fitted to the data. A multivariate Cox regression model was used to examine the survival association of CNA quartiles and age at diagnosis. The results indicated that all the variables examined are significantly associated with OS, while only CNA quartiles are significantly associated with DSS (Supplementary Table 18).

A univariate Cox analyses was then carried out which indicated that there was a significant association between DSS and CNA quartiles (Supplementary Table 16, Wald test *p*-value = 0.025). This is in agreement with the KM analysis (Figure 4B). PH assumptions were checked using statistical tests and graphical diagnostics based on scaled Schoenfeld residuals as the Cox PH model makes several assumptions. The test was not statistically significant for each of the variables, and the global test was also not statistically significant (Supplementary Figures 21-22). Hence, it can be concluded that the association between CNA quartiles and DSS in lumA grade 3 breast cancer patients in the METABRIC data was a direct result of CNA quartiles.

### Stratification of Luminal Cancers

Analyses carried out by Tishchenko et al. [11] on the transcriptomic and genomic landscape of luminal breast cancers in both the METABRIC and the Research Online Cancer Knowledgebase (ROCK) datasets suggested that the rigid stratification of luminal breast cancers into lumA and lumB intrinsic molecular subtypes is equivocal. These authors identified the top ten most up-regulated genes in all luminal samples and observed that they were primarily associated with cell proliferation (Supplementary Table 2).

Four quantiles comprising approximately 25% of METABRIC luminal patients were then defined based on the relative expression of these top ten genes. The progression from quantile 1 to quantile 4 showed an increase in patient risk level and an approximately continuous transition in the proportion of lumA and lumB subtypes (Tishchenko et al.’s Figure 5 [11]). The mixing between lumA and lumB classification begins in quantile 2 and reaches a peak of ambiguity in quantile 3 with some mixing still observed in quantile 4. The authors proposed that this reflects a continuous variation of a molecular profile with increasing genomic damage [11].

**Fig 5.**
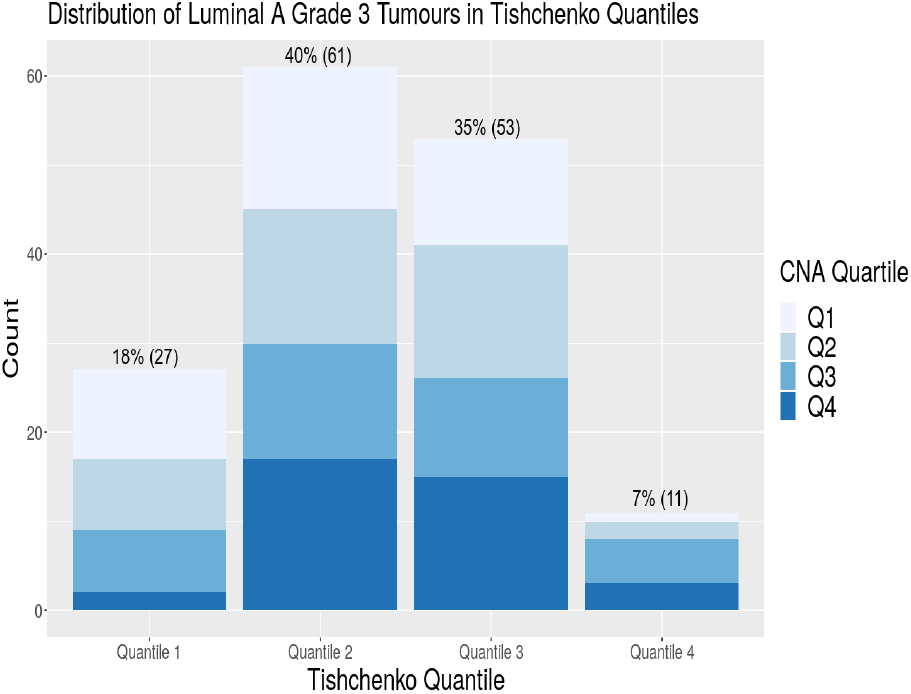
Distribution of CNA burden derived lumA grade 3 tumours corresponding to the original gene expression derived quantiles of Tishchenko et al. (2016).

Our CNA score correlates with the transcriptomic ranking of Tishchenko et al., as expected. Cross referencing revealed that of the 152 lumA grade 3 cases common to this study and the Tischchenko et al. [11] study, 40% and 35% were assigned by the latter authors to their quantiles 2 and 3 respectively (Figure 5). These results suggest that a large proportion of the lumA grade 3 cases have gene expression levels in transition between lumA and lumB subtype classification, and correspond to more at-risk lumA patients. Furthermore, the 37 lumA grade 3 cases with the highest CNA scores that we assigned to Q4 are largely found in Tishchenko et al. quantiles 2 and 3 (Supplementary Figure 32). This is consistent with increased GI detected by our CNA score preceding the transition in expression of cell proliferation genes. Together with the association of CNA score with survival outcome for lumA grade 3 cases, this suggests GI profiles could be informative for treatment decisions for luminal breast cancers.

## Discussion

We analysed the effect of CNA burden on breast cancer survival by implementing a series of statistically robust tools for interrogation of the rich and well annotated METABRIC dataset. We assigned each patient to a group by quartile segmentation of the summed distribution profile of absolute CNA scores, providing a first order measure of CNA burden that is a more realistic representation of GI compared to the more simplistic binary segmentation used in previous studies [1]. Our analysis revealed that the presence of high CNA levels in the tumour genomes of patients with lumA grade 3 breast cancer is associated with worse DSS outcomes, based on the clinical information and CNA profiles of 1175 luminal breast cancer patients registered in the METABRIC archive.

The observed difference in survival outcomes could be the result of either a true association between survival and CNA quartiles, or the result of confounding factors. LumA and lumB cases were considered separately since the PAM50 subtype was determined to be a significant confounding factor in the overall luminal analysis. A significant association was again observed between survival outcomes and CNA quartiles for both subtypes.

It was determined that NPI was one of the most significant confounding factors for the association in both lumA and lumB subtypes using the Kruskal-Wallis test. NPI incorporates tumour grade so lumA and lumB patients were further stratified based on grade. CNA score quartiles within lumA grade 1 and 2 and lumB grade 2 and 3 cases were not associated with OS or DSS whereas CNA quartiles within lumA grade 3 and lumB grade 1 cases did show a statistically significant association with DSS. Due to the small sample size per quartile for lumB grade 1 patients this association was viewed with caution. Further analysis of lumA grade 3 cases revealed that CNA score quartiles were associated with breast cancer prognosis independent of other strong clinical predictors.

Recent studies have proposed that lumA tumours may evolve into lumB tumours through the stochastic acquisitions of mutations in genes associated with worse prognosis [11, 14]. Tishchenko et al. (2016) used the top ten up-regulated genes in all METABRIC luminal cases to rank them by average expression and assign quantiles with approximately 25% of patients in each group. The incidence of lumB tumours was found to increase along with the patients risk level in the progression from quantile 1 to 4 by these authors. This led to their hypothesis that luminal tumours represent a continuum whose subtype range correlates with increasing genomic damage.

The lumA grade 3 tumours identified in our analysis largely correspond to quantiles 2 and 3 of the Tishchenko et al. study and occupy the region of PAM50 subtype stratification where the boundaries between lumA and lumB cases overlap. This implies that the lumA grade 3 cases we have identified display gene expression levels that more closely resemble those associated with the lumB category. Therefore, our work provides further support for the proposal of a gradient in luminal classification [11] by providing a robust statistical validation of the association between CNA burden and survival outcome for lumA cases at the boundary where lumA and lumB cases overlap in cell profileration gene expression.

LumA grade 3 patients who belong to both higher CNA score quartiles and higher gene expression quantiles are at particular risk for long term survival outcome. This has potential clinical utility because these patients are potentially not well stratified by the PAM50 subtype but can be identified by the simple measure of a CNA score. Therefore, patients classified as lumA grade 3 that display gene expression levels more akin to lumB tumours and also have high CNA burden may benefit from the more aggressive treatment regime used for lumB patients in contrast to standard endocrine therapy for lumA patients [30].

We defined CNA score as the sum of the absolute CNA values over all genes per patient then split the patients into quartiles based on their score. This CNA score definition enables unbiased analysis, maintains the easy interpretation of the data, and provides sufficient samples per CNA quartile to implement meaningful statistical analyses. However, this definition ignores the direction of copy number change and is acknowledged as a simplistic representation of the CNA landscape in tumour cells. Fine grained features including length of the CNA, whether it is an amplification or deletion, and the genomic location of the CNA are not considered. The analysis could potentially be made more sensitive by use of a richer metric for CNA score, although the smaller sample groups available following such fine grained segmentation of the METABRIC luminal cohort would likely compromise rigorous statistical analysis. Expanded datasets like METABRIC that combine high quality genomic and transcriptomic profiling with long-term clinical follow-up are required to provide sufficient cases for independent discovery and validation sets without such sample size concerns.

Overall, this work demonstrates a practical pathway towards the goals of personalised medicine, whereby a more individualised approach to classifying breast cancers can improve the success of treatment interventions by providing tailored therapeutic strategies based on the genomic profile of an individual tumour [31]. For example, a simple measure of CNA burden obtained from biopsy or resected tumour sample material could provide a prognostic biomarker to stratify a luminal breast cancer patient in addition to tumour grade and PAM50 subtype.

## Conclusion

It is important to identify features of luminal breast cancer that have prognostic value to aid in the identification of patients with a greater risk of lethal disease because the number of women diagnosed is increasing and the majority of cases belong to luminal subtypes. We analysed freely available clinical and genomic patient data from the METABRIC dataset to study the impact of CNA burden on overall survival within the luminal subtypes. We observed that CNA quartiles based on absolute CNA score are a prognostic factor for breast cancer survival outcomes in a subset of patients suffering from lumA grade 3 breast cancer. We further demonstrated that the lumA grade 3 cases in our study lie in the ambiguous region between lumA and lumB subtype classifications identified in an earlier analysis of gene expression levels from the same METABRIC patient samples. Women diagnosed with lumA grade 3 breast cancer that have gene expression levels more similar to lumB and who possess a CNA burden within our derived quartiles 3 or 4 may benefit from more aggressive therapy. This work progresses efforts to incorporate individual genomic landscapes into more nuanced classifications of breast cancer cases, with the goal of personalising therapeutic interventions to optimise long term survival outcomes for patients.

## Data Availability

All processed data products from the METABRIC consortium used in this work are freely available courtesy of the cBioportal for Cancer Genomics webportal.

https://www.cbioportal.org/study/summary?id=brca_metabric

## Supporting information

**S1 Fig. CNA quartiles within PAM50 subtypes in METABRIC luminal cases**

**S2 Fig. CNA quartiles within PAM50 subtypes in METABRIC luminal cases**

**S3 Fig. Assessing OS proportional hazard assumptions in METABRIC luminal cases**

**S4 Fig. Assessing DSS proportional hazard assumptions in METABRIC luminal cases**

**S5 Fig. Density plot of CNA score distribution in METABRIC lumA cases**

**S6 Fig. OS according to CNA score quartiles in METABRIC lumA cases**

**S7 Fig. DSS according to CNA score quartiles in METABRIC lumA cases**

**S7 Fig. Assessing OS proportional hazard assumptions in METABRIC lumA cases**

**S8 Fig. Assessing DSS proportional hazard assumptions in METABRIC lumA cases**

**S9 Fig. Density plot of CNA score distribution in METABRIC lumB cases**

**S10 Fig. OS according to CNA score quartiles in METABRIC lumB cases**

**S11 Fig. DSS according to CNA score quartiles in METABRIC lumB cases**

**S12 Fig. Assessing OS proportional hazard assumptions in METABRIC lumB cases**

**S13 Fig. Assessing DSS proportional hazard assumptions in METABRIC lumB cases**

**S14 Fig. Density plot of CNA score distribution in METABRIC lumA grade 1 cases**

**S15 Fig. OS according to CNA score quartiles in METABRIC lumA grade 1 cases**

**S16 Fig. DSS according to CNA score quartiles in METABRIC lumA grade 1 cases**

**S17 Fig. Density plot of CNA score distribution in METABRIC lumA grade 2 cases**

**S18 Fig. OS according to CNA score quartiles in METABRIC lumA grade 2 cases**

**S19 Fig. DSS according to CNA score quartiles in METABRIC lumA grade 2 cases**

**S20 Fig. Density plot of CNA score distribution in METABRIC lumA grade 3 cases**

**S21 Fig. Assessing OS proportional hazard assumptions in METABRIC lumA grade 3 cases**

**S22 Fig. Assessing DSS proportional hazard assumptions in METABRIC lumA grade 3 cases**

**S23 Fig. Density plot of CNA score distribution in METABRIC lumB grade 1 cases**

**S24 Fig. OS according to CNA score quartiles in METABRIC lumB grade 1 cases**

**S25 Fig. DSS according to CNA score quartiles in METABRIC lumB grade 1 cases**

**S26 Fig. Density plot of CNA score distribution in METABRIC lumB grade 2 cases**

**S27 Fig. OS according to CNA score quartiles in METABRIC lumB grade 2 cases**

**S28 Fig. DSS according to CNA score quartiles in METABRIC lumB grade 2 cases**

**S29 Fig. Density plot of CNA score distribution in METABRIC lumB grade 3 cases**

**S30 Fig. OS according to CNA score quartiles in METABRIC lumB grade 3 cases**

**S31 Fig. DSS according to CNA score quartiles in METABRIC lumB grade 3 cases**

**S32 Fig. Relationship of lumA grade 3 breast cancers in Q4 based on quantiles of Tishchenko (2016)**

**S1 Table. List and description of the clinical variables recorded within the METABRIC cohort**

**S2 Table. List of top ten most up-regulated genes in the Tishchenko et al. [1] study**

**S3 Table. Univariate Cox regression on clinical variables and CNA quartiles associated with METABRIC luminal cases**

**S4 Table. Assessment of association between CNA quartiles and clinical variables that showed a significant association with either OS, DSS or both in METABRIC luminal cases**

**S5 Table. Multivariate Cox regression on CNA quartiles and top three associated variables in METABRIC luminal cases**

**S6 Table. Univariate Cox regression on clinical variables and CNA quartiles associated with METABRIC lumA cases**

**S7 Table. Assessment of association between CNA quartiles and clinical variables that showed a significant association with either OS, DSS or both in METABRIC lumA cases**

**S8 Table. Multivariate Cox regression on CNA quartiles and top four associated variables in METABRIC lumA cases**

**S9 Table. Multivariate Cox regression on CNA quartiles and top three associated variables in METABRIC lumA cases**

**S10 Table. Multivariate Cox regression on CNA quartiles and top two associated variables in METABRIC lumA cases**

**S11 Table. Multivariate Cox regression on CNA quartiles and top associated variable in METABRIC lumA cases**

**S12 Table. Univariate Cox regression on clinical variables and CNA quartiles associated with METABRIC lumB cases**

**S13 Table. Assessment of association between CNA quartiles and clinical variables that showed a significant association with either OS, DSS or both in METABRIC lumB cases**

**S14 Table. Multivariate Cox regression on CNA quartiles and top four associated variables in METABRIC lumB cases**

**S15 Table. Multivariate Cox regression on CNA quartiles and top three associated variables in METABRIC lumB cases**

**S16 Table. Univariate Cox regression on clinical variables and CNA quartiles associated with METABRIC lumA grade 3 cases**

**S17 Table. Assessment of association between CNA quartiles and clinical variables that showed a significant association with either OS, DSS or both in METABRIC lumA grade 3 cases**

**S18 Table. Multivariate Cox regression on CNA quartiles and associated clinical variables in METABRIC lumA grade 3 cases**

**S19 Table. DSS Multivariate Cox regression on CNA quartiles in METABRIC lumA grade 3 cases**

**S1 File. Supplementary Materials** All figures, tables and additional results for this study

## Acknowledgments

The authors are grateful to careful reading of the manuscript by Dr. E. Holian and Dr. P. Ó Broin. This publication has emanated from research conducted with the financial support of Science Foundation Ireland under Grant number [18/CRT/6214].

